# Assessing the environmental impact of medicines in Italy using data from the Italian Medicines Agency

**DOI:** 10.1101/2024.05.16.24307469

**Authors:** Valentina Giunchi, Michele Fusaroli, Agnese Cangini, Filomena Fortinguerra, Simona Zito, Andrea Pierantozzi, Carlotta Lunghi, Elisabetta Poluzzi, Francesco Trotta

## Abstract

**Aim:** This study builds upon the environmental risk analysis presented in the 2022 National Report on Medicines Use in Italy by the Italian Medicines Agency and aims to assess the environmental risk posed by medicines in Italy and in its regions.

**Methods:** The analysis selected 90 medicines based on three criteria: high utilization, low PNEC, and inclusion or candidacy for the European Watch List. For each medicine, the environmental risk was computed as the ratio between the Predicted Environmental Concentration (PEC) and the Predicted No Effect Concentration (PNEC). PEC was derived following the approach of the Swedish Association of Pharmaceutical Industries and Italian drug utilization data. The risk was classified high if the ratio was greater than 10, and moderate if greater than 1.

**Results:** Overall, 13 medicines were identified as posing a high risk, including cardiovascular agents, antibiotics, analgesics, antidepressants, and antiparasitic agents. The high risk was driven by either a very low PNEC (e.g., estradiol and lacidipine), and high utilization (e.g., amoxicillin, ibuprofen, and diclofenac). Regional analysis showed higher risk due to high consumption for azithromycin and ofloxacin in Central and Southern Italy, and for levonorgestrel in Northern Italy.

**Conclusion:** This study points to the need of prioritizing targeted sampling in surface waters for medicines estimated at high risk. To prevent and mitigate the risk, a more conscious clinical practice coupled with appropriate waste management are required.

**What is already known about this subject:**

- Medicines represent a growing concern as a source of contamination in water systems.
- Traditional surface water samplings are resource-expensive and should be supported by estimation methods.
- The Italian Medicines Agency’s National report on medicines use included, for the first time in its 2022 version, an assessment of the environmental impact.

**What this study adds:**

- 90 medicines with either high utilization, low Predicted No-Effect Concentration, or present in the European Watch List were included.
- 13 medicines were at high environmental risk either because of their high toxicity to aquatic species or their high consumption in Italy.
- Regional differences potentially reflect various social and prescribing habits.

## Introduction

The use of medicines is one of the challenges posed by human activities to environmental sustainability^1^. After consumption, medicines are released into the environment through excretion in urine and feces, either unchanged or as inactivated or still as active metabolites. Additionally, they can be released directly in wastewater in the case of topically applied formulations or improper disposal of medications through toilets. This enables medicines to enter wastewater and, subsequently, surface waters, potentially causing adverse effects on the fauna and flora in these environments^1^. The exposure of aquatic organisms to medicines can result in various adverse reactions that threaten the ecosystem. For example, exposure to hormonal agents may lead to the feminization of male fish, certain analgesics can induce nephrotoxicity in various animal species, and the presence of antibiotics can contribute to the development of resistance in animals, plants, and humans^2–4^.

At the European level, measures have been undertaken to assess the presence and environmental risk associated with medicines. These measures have concentrated explicitly on monitoring surface waters, as they represent the first environment affected by the consequences of human medicine use. Since 2006 the European Medicines Agency (EMA) has required pharmaceutical manufacturers to include an Environmental Risk Assessment (ERA) as part of the European Public Assessment Report (EPAR) during the marketing authorization process for a pharmaceutical product. The ERA should provide information on the toxicity of the active pharmaceutical ingredients to aquatic organisms, along with details on the risk based on expected consumption^5^. Furthermore, in 2008 the European Commission (EC) introduced a compulsory monitoring system for surface water aimed at tracking a group of chemical substances, known as Watch List, which may pose harm to the environment. The monitoring campaign began in 2015, periodically reviewing the list of monitored substances, which include also human medicines^6^. In March 2019, the European Commission presented the “Strategic Approach to Pharmaceuticals in the Environment”, which includes actions aimed at countering the negative effects of medicines on the environment throughout their entire lifecycle, from design and production to use and disposal^7^.

However, the task of monitoring medicines in surface waters is a resource-intensive endeavor that is often limited to a specific set of substances. Therefore, it is essential to prioritize the substances for monitoring by employing estimation methods that go beyond the pre-marketing estimates supplied by manufacturers seeking marketing authorization from the EMA. A commonly adopted measure for estimating the environmental impact of medicines is the Risk Quotient (RQ), calculated as the ratio between the Predicted Environmental Concentration (PEC) and the Predicted No-Effect Concentration (PNEC)^5^. The PNEC represents a tolerability threshold for organisms exposed to the medicines derived from in-vivo tests^8^, while the PEC can be estimated in-silico using various approaches. Currently, calculating the PEC commonly relies on the method proposed by the Swedish Association of Pharmaceutical Industries (Lif - Läkemedelsindustriföreningen), which adapts the EMA’s approach for post-marketing scenarios^9,10^. Lif has incorporated an environmental risk section on the FASS (Farmacevtiska specialiteter i Sverige) website (www.fass.se) as part of this approach, providing information on each pharmaceutical product supplied in Sweden. Building on this initiative, the Region Stockholm has developed a dedicated web-based database for disseminating environmental information on medicines, including those found in FASS and other documents^11,12^. Inspired by the Swedish example, the Finnish Pharmaceutical Information Centre (Pharmaca) has integrated environmental information on medicines into the Pharmaca Fennica online service^13,14^. Other countries, such as Norway and Italy, have contributed to the development of their country-specific environmental risk assessments for medicines through publications in the scientific literature^15,16^.

The Italian Medicines Agency (AIFA – Agenzia Italiana del Farmaco) recently joined this initiative to map the environmental impact of medicines. In its 2022 annual OsMed report on national drug utilization (English version released in December 2023), a section was dedicated to estimate the environmental risk in Italy based on drug utilization data^17^.

This study aimed at assessing environmental risks of medicines used in Italy at the national, macro-area, and regional levels.

## Methods

### Selection of medicines

The environmental risk assessment was carried out for medicines that fulfilled the following criteria:

1. The first 30 medicines for human use most consumed in Italy in 2022.
2. Medicines for human use included or candidates in at least one version of the Watch List^18–21^.
3. Medicines for human use with the highest toxicity to aquatic animals and plants, based on the PNEC value of each active ingredient^22^. Some medicines meeting this criterion have been excluded from the analysis as they are not currently available on the Italian market (see **Table S1**).

### Drug Utilization sources

All Italian reimbursement categories, including over-the-counter (OTC) and hospital sales, were taken into account to ascertain the total sales volume of each selected medicine. Data were extracted from the Medicines Utilization Monitoring Centre (OsMed) database, which contains Information on the number of packages and related doses supplied in the 2022. The total kilograms for each active ingredient were estimated without considering the administration route or the ATC code.

### Risk computation

The environmental risk of medicines for surface waters was assessed by calculating the ratio between PEC and PNEC. It was computed for the whole Italian territory, the three main macro-areas of Italy (North, Center, and South and Islands), and each Italian region in 2022. Based on this assessment, environmental risk was classified as high when the PEC/PNEC ratio was greater than or equal to 10, moderate when between 1 and 10, low when between 0.1 and 1, and negligible when less than or equal to 0.1^10^.

The PEC of each pharmaceutical was computed using the Lif approach^10^:

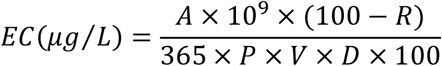

Specifically:

- *A* represents the total amount (in kilograms) of pharmaceutical supplied.
- *R* represents the pharmaceutical removal rate (%) through volatilization, hydrolysis, or biodegradation. Since specific information is not available, a value of 0% is assumed by default^8^.
- *P* represents the population size. P was calculated as the average between the residents on January 1^st^ 2022, and the residents on January 1^st^, 2023^23,24^.
- *V* represents the volume of daily per-capita wastewater production (L/day). It is by default set to 200 L/day according to the European Chemicals Agency (ECHA) proposal^8^.
- *D* represents the wastewater dilution factor produced by river flow and was set to 10, following the proposal of the ECHA^8^.

PNEC values were extracted from an open-access PNEC repository (https://osf.io/xtg8z/)^22,25^, which retrieved them from the NORMAN ecotoxicology database “lowest PNEC”^26^, the Watch List working documents^18–21^, the web-based database of the Region Stockholm “pharmaceuticals and environment”^12^ and, where needed, the scientific literature.

### Link with ATC classification

Medicines were categorized according to the WHO-ATC classification (2023 version)^27^. In cases where an active ingredient was associated with multiple ATC codes, preference was given to the most frequently used code.

## Results

A total of 90 medicines were identified based on our selection criteria and thus included in this analysis for environmental risk assessment (see **Table S2**). The majority of medicines belonged to cardiovascular agents (ATC C, n=21), anti-infectives for systemic use (ATC J, 16), and antineoplastic and immunomodulating agents (ATC L, 14). Six medicines, namely allopurinol, diclofenac, estradiol, ethinylestradiol, levonorgestrel, and metformin, were selected based on more than one criterion.

At the Italian level, 13 medicines were identified as posing a high risk to surface waters (see **Figure 1** and **Table S3**). Among them, two cardiovascular agents (ATC C, olmesartan and lacidipine) and four antibiotics (ATC J, rifaximin, ofloxacin, azithromycin, and amoxicillin). Other substances at high risk included the estrogen estradiol (ATC G), two nonsteroidal anti-inflammatory drugs (NSAIDs), such as ibuprofen and diclofenac (ATC M), two antidepressants—venlafaxine and sertraline (ATC N), the ectoparasiticide permethrin and the antiprotozoal atovaquone (ATC P). Furthermore, 23 medicines were categorized as having a moderate risk. The remaining 54 medicines were identified to have a low or insignificant risk according to assessments at the Italian level (see **Figure 1** and **Table S3**).

**Figure 1.**
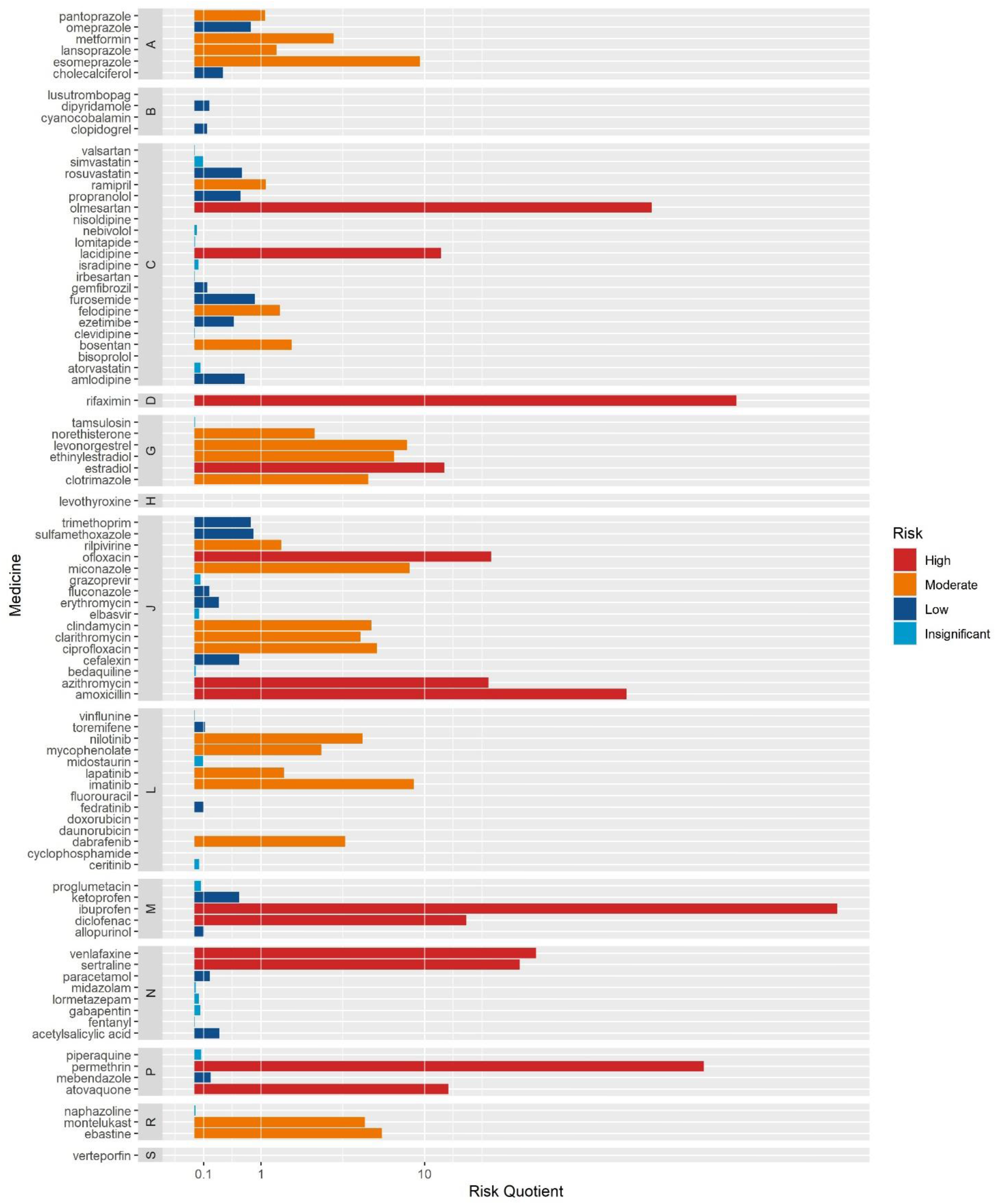
Risk level for the 90 selected medicines in Italy in 2022

The high risk stemmed from a very low PNEC for permethrin, estradiol, lacidipine, and atovaquone. Although permethrin and atovaquone are not frequently used, their high dose per quantity (2.25 grams) contributed to the risk profile. In contrast, estradiol and lacidipine are more commonly used, albeit with a lower quantity per dose. Conversely, other medicines at high risk, namely amoxicillin, azithromycin, ofloxacin, diclofenac, and ibuprofen have high PNEC values, but their widespread use and the high quantity per dose exacerbated their risk (see **Table S3**).

The analysis of the environmental risk at the macro-area level showed that risk was influenced by drug utilization patterns. Specifically, the high risk associated with lacidipine, ofloxacin, and azithromycin was driven by elevated consumption in the central and southern regions. Atovaquone’s high risk was uniquely influenced by the elevated consumption observed in Central Italy. Other notable patterns included the high risk of levonorgestrel restricted to Northern Italy, and the high risk of clotrimazole, clindamycin, ciprofloxacin, imatinib, montelukast, and ebastine restricted to Central Italy. Conversely, miconazole exhibited a high risk in both Central and Southern Italy (see **Figure 2**). These patterns are reflective of the distinctive drug utilization patterns within individual regions and the subsequent local environmental risks (see **Figure S1**).

**Figure 2.**
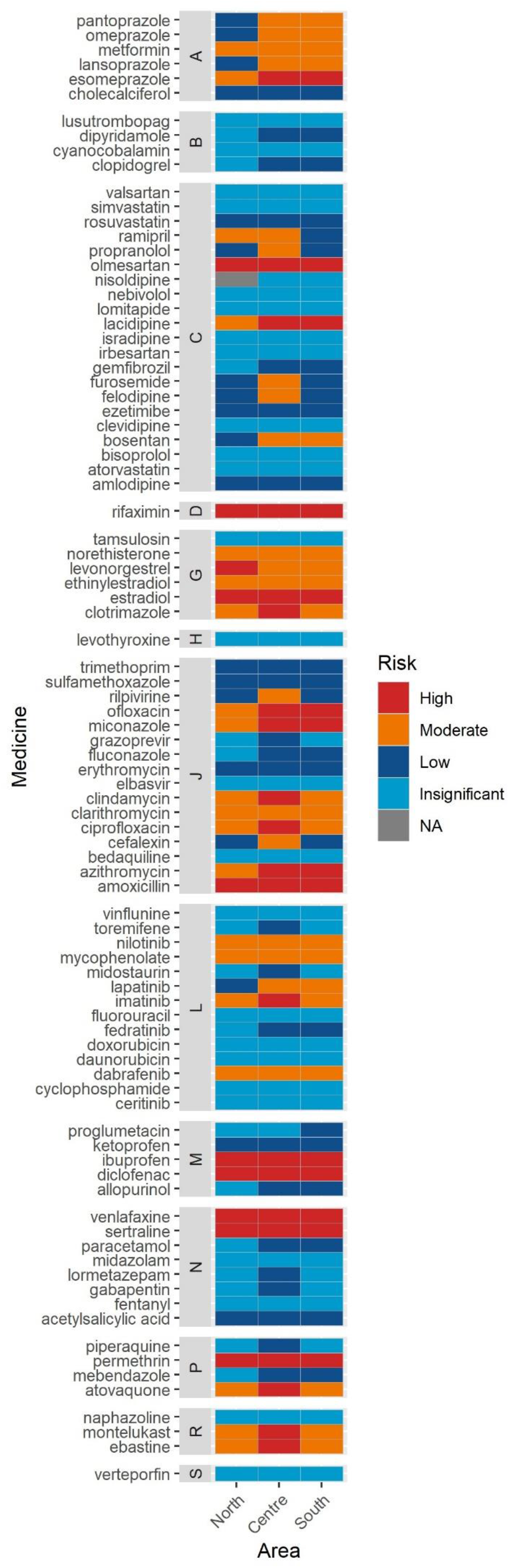
Risk level of the 90 selected medicines in 2022 divided by Italian macro-area

## Discussion

To our knowledge, this is the first study investigating the environmental impact of an extended range of medicines in Italy, starting from drug utilization data. We identified different high-risk medications, including NSAIDs, hormonal agents, antidepressants, cardiovascular drugs, and antibiotics.

The majority of therapeutic classes considered in this analysis exhibited at least one medicine posing either a high or moderate level of risk. This suggested that the environmental risk was widespread and distributed across various types of medicines. Notably, the only classes that did not show any risk in this analysis were the ATC B class of blood agents and the ATC S class of sensory organs agents. However, it’s important to acknowledge that the number of medicines from these classes included in the assessment was relatively limited and that most sensory agents have also another ATC code.

Commonly used medicines posing a high risk to Italian surface waters were the NSAIDs such as diclofenac and ibuprofen. The environmental impact of diclofenac has widely been studied and it is recognized to cause severe environmental damage^28,29^, with a high risk being acknowledged in different countries^30^. On the other hand, ibuprofen, despite its high risk, driven essentially by wide consumption, may have an overall lower environmental impact because it had a lower potential for bioaccumulation and persistence^30,31^. Otherwise, ketoprofen, acetylsalicylic acid, and paracetamol, belonging to the same therapeutic area, showed a low risk. Despite their similarly high consumption, their PNEC values were higher compared to those of diclofenac and ibuprofen, resulting in a lower environmental risk.

Hormonal agents commonly employed for contraceptive purposes, and to lesser extent for menopausal symptom treatment, were associated with either a high or moderate level of risk. These substances are widely acknowledged for their potential to cause harm when present in surface water because of their capability to disrupt the hormonal equilibrium of different aquatic species. For example, they can cause the feminization of male fish and consequently disrupt ecological equilibrium^2^. Contraceptives represented a striking example of medicines with low opportunity to reduce the uses to limit the environmental impact. Rather, the only measure to limit it would be to improve wastewater treatment systems.

Another pharmaceutical compound identified as posing a high risk to the environment is permethrin, an antiparasitic agent commonly employed for the treatment and prevention of head lice and scabies^32^. Since permethrin is extensively utilized in agriculture as a broad-spectrum insecticide, the hazard to the environment is probably even higher^33,34^. Also, the antiprotozoal atovaquone has been identified as posing a high environmental risk. While these latter two medicines have been identified as potential high-risk emerging contaminants on the basis of PNEC values obtained in experimental conditions, their effects on real aquatic ecosystems have not been thoroughly studied^35–37^.

The antidepressants sertraline and venlafaxine, respectively a selective serotonin reuptake inhibitor (SSRI) and a serotonin-norepinephrine reuptake inhibitor (SNRI), were identified as posing a high environmental risk. They are widely used to treat a broad spectrum of prevalent psychiatric disorders, including major depression and anxiety disorders^38^. The primary toxic effect observed in exposed fish was alteration in behavior, although there are conflicting findings^39–42^. Clinical alternatives, such as citalopram and escitalopram, have higher PNEC^22^, potentially rendering their environmental risk lower.

Among the ATC class C medicines, olmesartan, an angiotensin II receptor blocker^43^, and lacidipine, a calcium channel blocker also used for hypertension^44^, were estimated to pose a high environmental risk. In contrast, the angiotensin II receptor blocker, valsartan, resulted in an insignificant impact on the environment. Among the calcium channel blockers class, felodipine presented a moderate risk, while amlodipine and nisoldipine presented a low risk. Moreover, ramipril, an ACE inhibitor, was estimated to pose a moderate risk, while the beta-blockers bisoprolol, nebivolol, and propranolol resulted in a low or insignificant environmental risk. Certain studies emphasized that medicines within the same therapeutic class may exhibit distinct behaviors during wastewater treatment. For instance, among angiotensin II receptor blockers, olmesartan degrades more slowly than valsartan, adding complexity to their risk assessment^45^. A few studies have identified adverse reactions, including reproductive problems and issues related to biochemistry, such as disturbances in lipid metabolism, oxidative stress, and steroid levels^46^, but, overall, the effects of cardiovascular agents on aquatic organisms and plants remained insufficiently explored. Given the vast array of medicines available for the treatment of hypertension and the large number of people affected, it is crucial to consider the environmental impact in the overall assessment of these medications even at the moment of medical prescription. This should be included in a comprehensive assessment of their sustainability, along with their clinical risk-benefits, costs, and accessibility to patients.

Finally, a high environmental risk was estimated for certain antibacterials and antibiotics. The penicillin amoxicillin, the macrolide azithromycin, and the fluoroquinolone ofloxacin were identified as high-risk medicines. Rifaximin, an antibiotic mainly used to treat traveler’s diarrhea caused by *Escherichia Coli*, was also classified as high-risk. However, it is important to note that it could be purchased in Italy for use during travels abroad. Additionally, this medicine may be overused for the treatment of certain diseases, such as diverticulitis^47^. Moderate risk was estimated for ciprofloxacin (a fluoroquinolone) and clarithromycin (a macrolide), while erythromycin, another macrolide, was estimated at low risk. The elevated risk associated with commonly used antibacterials in Italy raised significant concerns, particularly regarding potential adverse impacts on aquatic flora and fauna^48^. Beyond posing immediate threats to the aquatic environment, these antibacterials also exhibit a worrisome capacity for bioaccumulation, further amplifying the risk. Moreover, the potential for the development of antibiotic-resistant strains adds a critical layer to the potential consequences, emphasizing the need for careful monitoring and management of antibiotic use to safeguard both environmental and public health^4,49,50^. Similarly, to cardiovascular drug classes, the environmental impact of each antimicrobial agent should be added to the clinical profile to provide physicians with additional information to make the appropriate therapeutic choice.

We identified some differences in the environmental risk of certain medicines across Italian regions. These differences reflected local drug prescription attitudes and consumption patterns and may reflect different healthcare practices and socio-economic disparities. Regions with higher consumption rates of high-risk medicines may benefit from targeted initiatives aimed at both physicians and patients, promoting responsible drug prescription practices, raising awareness about the environmental consequences of improper drug disposal and implementation of advanced wastewater treatment systems.

### Limitations and future perspectives

OsMed data, with their extensive spatial coverage at national, macro-area, and regional levels, along with their comprehensive coverage of all medicines supplied, offer an exhaustive picture of drug utilization in Italy and its associated risk on surface waters. This analysis may be useful for underlying the differences between the geographic areas and for the establishment of a routine of environmental risk estimation.

The primary limitation of this study is the utilization of default values for both the volume of wastewater produced and the removal rate at wastewater treatment plants in the risk derivation approach employed. Further, this analysis assumes a worst-case scenario where all consumed medicines are hypothesized to end up in surface waters. Elements such as human detoxification, correct disposal, and wastewater treatment, which have the potential to reduce surface water concentrations of medicines, were not included. Additionally, the computed PEC only accounted for human consumption, overlooking the potential contribution of veterinary medicines. Although the impact of veterinary medicines on surface water is often indirect, primarily through soil contamination, it could be significant for certain therapeutic classes such as antibiotics and hormones. To enhance the accuracy of future assessments, efforts should be directed toward collecting this data at the Italian, macro-area, and regional levels. Furthermore, we used both in-vivo and in-silico PNEC values without making a distinction between them. Specifically, in-silico PNEC values, derived through a Quantitative Structure-Activity Relationship (QSAR) approach, were found to be the lowest for 28 out of the 90 medicines analyzed, with 12 medicines having no other values available. All in-silico PNEC values were sourced from the NORMAN database. Finally, we shouldn’t solely focus on substances exceeding their PNEC. This is because several substances within the same class might share a toxic mechanism, potentially causing an effect when present together, even if each substance individually remained below the threshold of no effect. Additionally, different substances could exhibit a synergistic effect by acting at various points along the same pathway, or they might combine to form new substances with different mechanisms or potencies^51,52^. Further, low concentrations of medicines in the environment have the potential for bioaccumulation, subsequently leading to higher concentrations along the food chain – a phenomenon known as biomagnification. As a result, even medicines initially deemed low risk could impact secondary consumers^53^.

Proactive measures are essential for managing the high environmental risk associated with certain medicines, and the appropriate actions may vary depending on the therapeutic class. For example, in classes such as NSAIDs and antibiotics, limiting overconsumption, overprescription, and unnecessary self-treatment can lead to improvements in drug utilization. However, for medicines in classes like hormonal agents, where drug utilization may not be easily modifiable, it is crucial to take post-utilization measures. This may include the implementation of filtering systems in wastewater treatment plants to mitigate the impact on the environment^54,55^.

## Supporting information

script_data

## Acknowledgements

Part of the content was published in the OsMed 2022 report on medicines use in Italy (accessible at https://www.aifa.gov.it/en/-/l-uso-dei-farmaci-in-italia-rapporto-osmed-2022).

VG and CL were supported by Italian PON (Programma Operativo Nazionale) funds on green research. EP was supported by institutional research funds (Ricerca Fondamentale Orientata). MF is supported by funds for the PhD of the University of Bologna.

## Data availability statement

Data and scripts used in this analysis are available in the supplementary material.

## Author contribution

VG, MF, EP, CL, AC, FF, and SZ conceptualized and designed the study. VG, MF, CL, EP, AC, FF, SZ, AP curated drug utilization data. VG retrieved data other than drug utilization, and performed analyses and graphical visualization. VG wrote the original draft. MF, CL, EP, AC, FF, SZ revised and integrated the original draft. All the authors read and approved the final version.

