## supplementary for "Assessing the environmental impact of medicines in Italy using data from the Italian Medicines Agency"

**Supplementary material**

**Table S1** – Medicines excluded from lowest PNEC selection

| Pharmaceutical | Reason |
|---|---|
| Samatasvir | Still in phase II |
| Anacetrapib | Development abandoned in 2017 |
| Avasimibe | Used only in trials |
| Implitapide | Still in phase II |
| Bizelesin | Still in phase I |
| Vedroprevir | Still in phase II |
| Azelnidipine | Sold in Japan and India only |
| Triclocarban | Theoretically not used nowadays |
| Sovaprevir | Still in phase II |
| Efonidipine | Sold in Japan and India only |
| Cenicriviroc | Still in phase III |
| Linsitinib | Still in phase III |
| Rebastinib | Still in phase II |

**Table S2** – Medicines selected for the analysis

| ATC I Class | ATC code | Substance | Inclusion criteria |
|---|---|---|---|
| A | A02BC01 | omeprazole | Drug utilization |
| | A02BC02 | pantoprazole | Drug utilization |
| | A02BC03 | lansoprazole | Drug utilization |
| | A02BC05 | esomeprazole | Drug utilization |
| | A10BA02 | metformin | Drug utilization;Watch List |
| | A11CC05 | cholecalciferol | Drug utilization |
| B | B01AC04 | clopidogrel | Drug utilization |
| | B01AC07 | dipyridamole | Watch List |
| | B02BX07 | lusutrombopag | PNEC |
| | B03BA01 | cyanocobalamin | Drug utilization |
| C | C02KX01 | bosentan | PNEC |
| | C03CA01 | furosemide | Drug utilization |
| | C07AA05 | propranolol | Watch List |
| | C07AB07 | bisoprolol | Drug utilization |
| | C07AB12 | nebivolol | Drug utilization |
| | C08CA01 | amlodipine | Drug utilization |
| | C08CA02 | felodipine | PNEC |
| | C08CA03 | isradipine | PNEC |
| | C08CA07 | nisoldipine | PNEC |
| | C08CA09 | lacidipine | PNEC |
| | C08CA16 | clevidipine | PNEC |
| | C09AA05 | ramipril | Drug utilization |
| | C09CA03 | valsartan | Drug utilization |
| | C09CA04 | irbesartan | Watch List |
| | C09CA08 | olmesartan | Drug utilization |
| | C10AA01 | simvastatin | Drug utilization |

| | | | |
|---|---|---|---|
| | C10AA05 | atorvastatin | Drug utilization |
| | C10AA07 | rosuvastatin | Drug utilization |
| | C10AB04 | gemfibrozil | Watch List |
| | C10AX09 | ezetimibe | Drug utilization |
| | C10AX12 | lomitapide | PNEC |
| D | D06AX11 | rifaximin | PNEC |
| G | G01AF02 | clotrimazole | Watch List |
| | G03AC01 | norethisterone | Watch List |
| | G03AC03 | levonorgestrel | PNEC;Watch List |
| | G03CA01 | ethinylestradiol | PNEC;Watch List |
| | G03CA03 | estradiol | PNEC;Watch List |
| | G04CA02 | tamsulosin | Drug utilization |
| H | H03AA01 | levothyroxine | Drug utilization |
| J | J01CA04 | amoxicillin | Watch List |
| | J01DB01 | cefalexin | Watch List |
| | J01EA01 | trimethoprim | Watch List |
| | J01EC01 | sulfamethoxazole | Watch List |
| | J01FA01 | erythromycin | Watch List |
| | J01FA09 | clarithromycin | Watch List |
| | J01FA10 | azithromycin | Watch List |
| | J01FF01 | clindamycin | Watch List |
| | J01MA01 | ofloxacin | Watch List |
| | J01MA02 | ciprofloxacin | Watch List |
| | J02AB01 | miconazole | Watch List |
| | J02AC01 | fluconazole | Watch List |
| | J04AK05 | bedaquiline | PNEC |
| | J05AG05 | rilpivirine | PNEC |
| | J05AP10 | elbasvir | PNEC |
| | J05AP11 | grazoprevir | PNEC |
| L | L01AA01 | cyclophosphamide | Watch List |
| | L01BC02 | fluorouracil | Watch List |
| | L01CA05 | vinflunine | PNEC |
| | L01DB01 | doxorubicin | Watch List |
| | L01DB02 | daunorubicin | Watch List |
| | L01EA01 | imatinib | PNEC |
| | L01EA03 | nilotinib | PNEC |
| | L01EC02 | dabrafenib | PNEC |
| | L01ED02 | ceritinib | PNEC |
| | L01EH01 | lapatinib | PNEC |
| | L01EJ02 | fedratinib | PNEC |
| | L01EX10 | midostaurin | PNEC |
| | L02BA02 | toremifene | PNEC |
| | L04AA06 | mycophenolate | Watch List |
| M | M01AB05 | diclofenac | Drug utilization;Watch List |
| | M01AB14 | proglumetacin | PNEC |
| | M01AE01 | ibuprofen | Drug utilization |
| | M01AE03 | ketoprofen | Drug utilization |

| | M04AA01 | allopurinol | Drug utilization;Watch List |
|---|---|---|---|
| N | N01AH01 | fentanyl | Watch List |
| | N02BA01 | acetylsalicylic acid | Drug utilization |
| | N02BE01 | paracetamol | Drug utilization |
| | N02BF01 | gabapentin | Watch List |
| | N05CD06 | lormetazepam | Drug utilization |
| | N05CD08 | midazolam | Watch List |
| | N06AB06 | sertraline | Drug utilization |
| | N06AX16 | venlafaxine | Watch List |
| P | P01AX06 | atovaquone | PNEC |
| | P01BF05 | piperaquine | Watch List |
| | P02CA01 | mebendazole | Watch List |
| | P03AC04 | permethrin | PNEC |
| R | R01AA08 | naphazoline | Drug utilization |
| | R03DC03 | montelukast | PNEC |
| | R06AX22 | ebastine | PNEC |
| S | S01LA01 | verteporfin | PNEC |

**Table S3** – Risk quotients, risk classes, PEC, PNEC, and total drug utilization in kilograms for the 90 selected medicines in Italy, 2022

| Substance | DU (kg) | PEC | PNEC (µg/L) | RQ | Risk |
|---|---|---|---|---|---|
| acetylsalicylic acid | 227292.233 | 5.285 | 18.000 | 0.294 | Low |
| allopurinol | 93013.978 | 2.163 | 20.558 | 0.105 | Low |
| amlodipine | 6819.880 | 0.156 | 0.230 | 0.689 | Low |
| amoxicillin | 300471.054 | 6.986 | 0.078 | 89.566 | High |
| atorvastatin | 24272.278 | 0.564 | 8.500 | 0.066 | Insignificant |
| atovaquone | 466.674 | 0.011 | 0.001 | 13.073 | High |
| azithromycin | 16654.818 | 0.387 | 0.019 | 20.381 | High |
| bedaquiline | 1.485 | 0.000 | 0.003 | 0.013 | Insignificant |
| bisoprolol | 3091.188 | 0.072 | 92.000 | 0.001 | Insignificant |
| bosentan | 204.196 | 0.005 | 0.003 | 1.752 | Moderate |
| cefalexin | 2026.775 | 0.047 | 0.080 | 0.589 | Low |
| ceritinib | 2.403 | 0.000 | 0.001 | 0.049 | Insignificant |
| cholecalciferol | 205.671 | 0.005 | 0.014 | 0.347 | Low |
| ciprofloxacin | 21755.377 | 0.506 | 0.089 | 5.683 | Moderate |
| clarithromycin | 23978.310 | 0.558 | 0.120 | 4.646 | Moderate |
| clevidipine | 0.141 | 0.000 | 0.001 | 0.003 | Insignificant |
| clindamycin | 10072.303 | 0.234 | 0.044 | 5.322 | Moderate |
| clopidogrel | 19509.894 | 0.454 | 3.214 | 0.141 | Low |
| clotrimazole | 4396.261 | 0.102 | 0.020 | 5.111 | Moderate |
| cyanocobalamin | 42.090 | 0.001 | 33608.828 | 0.000 | Insignificant |
| cyclophosphamide | 176.731 | 0.004 | 6.964 | 0.001 | Insignificant |
| dabrafenib | 195.258 | 0.005 | 0.001 | 3.815 | Moderate |
| daunorubicin | 0.460 | 0.000 | 0.218 | 0.000 | Insignificant |
| diclofenac | 34473.326 | 0.802 | 0.050 | 16.031 | High |
| dipyridamole | 38.734 | 0.001 | 0.005 | 0.169 | Low |

| | | | | | |
|---|---|---|---|---|---|
| doxorubicin | 5.982 | 0.000 | 0.170 | 0.001 | Insignificant |
| ebastine | 726.463 | 0.017 | 0.003 | 6.054 | Moderate |
| elbasvir | 1.319 | 0.000 | 0.001 | 0.050 | Insignificant |
| erythromycin | 2496.319 | 0.058 | 0.200 | 0.290 | Low |
| esomeprazole | 12250.152 | 0.285 | 0.030 | 9.500 | Moderate |
| estradiol | 214.790 | 0.005 | 0.000 | 12.485 | High |
| ethinylestradiol | 10.570 | 0.000 | 0.000 | 7.022 | Moderate |
| ezetimibe | 3556.030 | 0.083 | 0.163 | 0.507 | Low |
| fedratinib | 7.860 | 0.000 | 0.002 | 0.101 | Low |
| felodipine | 96.974 | 0.002 | 0.002 | 1.436 | Moderate |
| fentanyl | 19.350 | 0.000 | 0.171 | 0.003 | Insignificant |
| fluconazole | 1809.268 | 0.042 | 0.250 | 0.168 | Low |
| fluorouracil | 704.824 | 0.016 | 58.500 | 0.000 | Insignificant |
| furosemide | 26755.784 | 0.622 | 0.707 | 0.880 | Low |
| gabapentin | 25708.471 | 0.598 | 10.000 | 0.060 | Insignificant |
| gemfibrozil | 3107.124 | 0.072 | 0.500 | 0.144 | Low |
| grazoprevir | 2.638 | 0.000 | 0.001 | 0.065 | Insignificant |
| ibuprofen | 384053.104 | 8.930 | 0.011 | 811.775 | High |
| imatinib | 958.006 | 0.022 | 0.003 | 8.839 | Moderate |
| irbesartan | 42789.243 | 0.995 | 700.000 | 0.001 | Insignificant |
| isradipine | 2.546 | 0.000 | 0.001 | 0.042 | Insignificant |
| ketoprofen | 53280.932 | 1.239 | 2.096 | 0.591 | Low |
| lacidipine | 254.206 | 0.006 | 0.000 | 12.062 | High |
| lansoprazole | 11143.394 | 0.259 | 0.192 | 1.349 | Moderate |
| lapatinib | 80.927 | 0.002 | 0.001 | 1.542 | Moderate |
| levonorgestrel | 5.615 | 0.000 | 0.000 | 8.160 | Moderate |
| levothyroxine | 102.583 | 0.002 | 12.000 | 0.000 | Insignificant |
| lomitapide | 0.346 | 0.000 | 0.001 | 0.006 | Insignificant |
| lormetazepam | 340.753 | 0.008 | 0.166 | 0.048 | Insignificant |
| lusutrombopag | 0.004 | 0.000 | 0.002 | 0.000 | Insignificant |
| mebendazole | 692.648 | 0.016 | 0.088 | 0.183 | Low |
| metformin | 1404001.809 | 32.644 | 10.000 | 3.264 | Moderate |
| miconazole | 9116.543 | 0.212 | 0.025 | 8.405 | Moderate |
| midazolam | 61.901 | 0.001 | 0.115 | 0.013 | Insignificant |
| midostaurin | 3.191 | 0.000 | 0.001 | 0.096 | Insignificant |
| montelukast | 466.593 | 0.011 | 0.002 | 4.909 | Moderate |
| mycophenolate | 15605.201 | 0.363 | 0.132 | 2.749 | Moderate |
| naphazoline | 126.893 | 0.003 | 0.370 | 0.008 | Insignificant |
| nebivolol | 1896.279 | 0.044 | 1.838 | 0.024 | Insignificant |
| nilotinib | 303.610 | 0.007 | 0.001 | 4.770 | Moderate |
| nisoldipine | 0.000 | 0.000 | 0.002 | 0.000 | Insignificant |
| norethisterone | 53.729 | 0.001 | 0.001 | 2.498 | Moderate |
| ofloxacin | 23554.914 | 0.548 | 0.026 | 21.064 | High |
| olmesartan | 20167.312 | 0.469 | 0.004 | 116.643 | High |
| omeprazole | 9615.485 | 0.224 | 0.280 | 0.798 | Low |
| pantoprazole | 31952.477 | 0.743 | 0.681 | 1.091 | Moderate |
| paracetamol | 986193.786 | 22.930 | 134.000 | 0.171 | Low |

| | | | | | |
|---|---|---|---|---|---|
| permethrin | 1728.905 | 0.040 | 0.000 | 200.992 | High |
| piperaquine | 3.318 | 0.000 | 0.001 | 0.073 | Insignificant |
| proglumetacin | 4.932 | 0.000 | 0.002 | 0.067 | Insignificant |
| propranolol | 2662.680 | 0.062 | 0.100 | 0.619 | Low |
| ramipril | 5193.455 | 0.121 | 0.110 | 1.102 | Moderate |
| rifaximin | 30199.349 | 0.702 | 0.002 | 283.128 | High |
| rilpivirine | 160.280 | 0.004 | 0.003 | 1.467 | Moderate |
| rosuvastatin | 7417.417 | 0.172 | 0.269 | 0.640 | Low |
| sertraline | 11597.071 | 0.270 | 0.009 | 28.685 | High |
| simvastatin | 10659.184 | 0.248 | 2.630 | 0.094 | Insignificant |
| sulfamethoxazole | 3662.083 | 0.085 | 0.100 | 0.851 | Low |
| tamsulosin | 104.986 | 0.002 | 0.346 | 0.007 | Insignificant |
| toremifene | 1.503 | 0.000 | 0.000 | 0.116 | Low |
| trimethoprim | 17064.212 | 0.397 | 0.500 | 0.794 | Low |
| valsartan | 44002.931 | 1.023 | 560.000 | 0.002 | Insignificant |
| venlafaxine | 8961.491 | 0.208 | 0.006 | 34.158 | High |
| verteporfin | 0.013 | 0.000 | 0.002 | 0.000 | Insignificant |
| vinflunine | 0.170 | 0.000 | 0.001 | 0.004 | Insignificant |

**Table S4** - Risk quotients, PEC (expressed in μg/L), and total drug utilization (expressed in kgs) for the 90 selected medicines in 2022 in each Italian macro-area. PNEC are the same as reported in Table S2, and risk can be interpreted as high when ≥10, moderate when ≥1, low when ≥0.1, insignificant otherwise.

| Substance | DU North | DU Centre | DU South | PEC North | PEC Centre | PEC South | RQ North | RQCentre | RQ South |
|---|---|---|---|---|---|---|---|---|---|
| acetylsalicylic acid | 107474.727 | 44373.528 | 75443.978 | 5.377 | 5.188 | 5.214 | 0.299 | 0.288 | 0.29 |
| allopurinol | 21114.803 | 39746.685 | 32152.49 | 1.056 | 4.647 | 2.222 | 0.051 | 0.226 | 0.108 |
| amlodipine | 3163.211 | 1466.741 | 2189.928 | 0.158 | 0.171 | 0.151 | 0.688 | 0.746 | 0.658 |
| amoxicillin | 61041.16 | 136030.433 | 103399.461 | 3.054 | 15.904 | 7.146 | 39.154 | 203.892 | 91.621 |
| atorvastatin | 11075.473 | 4617.659 | 8579.146 | 0.554 | 0.54 | 0.593 | 0.065 | 0.064 | 0.07 |
| atovaquone | 81.805 | 329.602 | 55.267 | 0.004 | 0.039 | 0.004 | 4.931 | 46.427 | 4.602 |
| azithromycin | 3520.36 | 6605.954 | 6528.503 | 0.176 | 0.772 | 0.451 | 9.27 | 40.648 | 23.748 |
| bedaquiline | 0.47 | 0.827 | 0.188 | 0 | 0 | 0 | 0.009 | 0.035 | 0.005 |
| bisoprolol | 586.458 | 1471.737 | 1032.993 | 0.029 | 0.172 | 0.071 | 0 | 0.002 | 0.001 |
| bosentan | 43.51 | 80.033 | 80.653 | 0.002 | 0.009 | 0.006 | 0.803 | 3.453 | 2.057 |
| cefalexin | 498.261 | 930.899 | 597.615 | 0.025 | 0.109 | 0.041 | 0.312 | 1.36 | 0.516 |
| ceritinib | 0.986 | 0.878 | 0.54 | 0 | 0 | 0 | 0.043 | 0.089 | 0.032 |
| cholecalciferol | 120.305 | 39.541 | 45.825 | 0.006 | 0.005 | 0.003 | 0.437 | 0.336 | 0.230 |
| ciprofloxacin | 4791.997 | 7790.211 | 9173.168 | 0.24 | 0.911 | 0.634 | 2.694 | 10.233 | 7.124 |
| clarithromycin | 4987.696 | 7865.095 | 11125.52 | 0.25 | 0.92 | 0.769 | 2.08 | 7.663 | 6.408 |
| clevidipine | 0.046 | 0.056 | 0.039 | 0 | 0 | 0 | 0.002 | 0.006 | 0.003 |
| clindamycin | 2059.145 | 4973.32 | 3039.839 | 0.103 | 0.581 | 0.21 | 2.341 | 13.215 | 4.775 |
| clopidogrel | 4956.125 | 7614.789 | 6938.98 | 0.248 | 0.89 | 0.48 | 0.077 | 0.277 | 0.149 |
| clotrimazole | 979.85 | 2041.408 | 1375.003 | 0.049 | 0.239 | 0.095 | 2.451 | 11.933 | 4.752 |
| cyanocobalamin | 19.55 | 8.729 | 13.811 | 0.001 | 0.001 | 0.001 | 0 | 0 | 0 |
| cyclophosphamide | 41.756 | 84.711 | 50.264 | 0.002 | 0.01 | 0.003 | 0 | 0.001 | 0 |
| dabrafenib | 40.038 | 98.28 | 56.94 | 0.002 | 0.011 | 0.004 | 1.683 | 9.656 | 3.307 |
| daunorubicin | 0.1 | 0.204 | 0.156 | 0 | 0 | 0 | 0 | 0 | 0 |
| diclofenac | 15482.158 | 6724.279 | 12266.889 | 0.775 | 0.786 | 0.848 | 15.492 | 15.723 | 15.957 |
| dipyridamole | 9.335 | 19.902 | 9.497 | 0 | 0.002 | 0.001 | 0.087 | 0.436 | 0.123 |
| doxorubicin | 1.116 | 2.969 | 1.897 | 0 | 0 | 0 | 0 | 0.002 | 0.001 |
| ebastine | 133.944 | 314.322 | 278.198 | 0.007 | 0.037 | 0.019 | 2.402 | 13.171 | 6.892 |
| elbasvir | 0.143 | 0.521 | 0.655 | 0 | 0 | 0 | 0.012 | 0.099 | 0.073 |

| | | | | | | | | | |
|---|---|---|---|---|---|---|---|---|---|
| erythromycin | 587.104 | 1032.674 | 876.54 | 0.029 | 0.121 | 0.061 | 0.147 | 0.604 | 0.303 |
| esomeprazole | 1952.242 | 5848.504 | 4449.407 | 0.098 | 0.684 | 0.308 | 3.258 | 22.806 | 10.257 |
| estradiol | 339.660 | 106.464 | 132.344 | 0.017 | 0.012 | 0.009 | 42.485 | 31.117 | 22.867 |
| ethinylestradiol | 6.125 | 1.927 | 2.602 | 0 | 0 | 0 | 8.756 | 6.436 | 5.139 |
| ezetimibe | 1611.726 | 748.697 | 1195.607 | 0.081 | 0.088 | 0.083 | 0.495 | 0.537 | 0.507 |
| fedratinib | 0.648 | 4.452 | 2.76 | 0 | 0.001 | 0 | 0.018 | 0.286 | 0.105 |
| felodipine | 18.03 | 57.616 | 21.328 | 0.001 | 0.007 | 0.001 | 0.575 | 4.29 | 0.939 |
| fentanyl | 11.126 | 3.129 | 5.096 | 0.001 | 0.000 | 0.000 | 0.003 | 0.002 | 0.002 |
| fluconazole | 337.143 | 682.294 | 789.831 | 0.017 | 0.08 | 0.055 | 0.067 | 0.319 | 0.218 |
| fluorouracil | 174.182 | 269.215 | 261.426 | 0.009 | 0.031 | 0.018 | 0 | 0.001 | 0 |
| furosemide | 5720 | 12608.467 | 8427.317 | 0.286 | 1.474 | 0.582 | 0.405 | 2.086 | 0.824 |
| gabapentin | 8193.999 | 11892.598 | 5621.874 | 0.41 | 1.39 | 0.389 | 0.041 | 0.139 | 0.039 |
| gemfibrozil | 799.308 | 1423.278 | 884.538 | 0.04 | 0.166 | 0.061 | 0.08 | 0.333 | 0.122 |
| grazoprevir | 0.286 | 1.042 | 1.31 | 0 | 0 | 0 | 0.015 | 0.129 | 0.096 |
| ibuprofen | 185815.682 | 81193.76 | 117043.661 | 9.297 | 9.493 | 8.089 | 845.157 | 862.956 | 735.407 |
| imatinib | 205.774 | 455.82 | 296.412 | 0.01 | 0.053 | 0.02 | 4.085 | 21.147 | 8.13 |
| irbesartan | 10123.216 | 16597.955 | 16068.072 | 0.506 | 1.94 | 1.111 | 0.001 | 0.003 | 0.002 |
| isradipine | 0.439 | 0.977 | 1.13 | 0 | 0 | 0 | 0.016 | 0.082 | 0.056 |
| ketoprofen | 23736.918 | 10429.351 | 19114.663 | 1.188 | 1.219 | 1.321 | 0.567 | 0.582 | 0.630 |
| lacidipine | 55.502 | 115.655 | 83.049 | 0.003 | 0.014 | 0.006 | 5.667 | 27.595 | 11.714 |
| lansoprazole | 2059.518 | 5467.012 | 3616.864 | 0.103 | 0.639 | 0.25 | 0.537 | 3.329 | 1.302 |
| lapatinib | 13.748 | 28.217 | 38.962 | 0.001 | 0.003 | 0.003 | 0.564 | 2.704 | 2.207 |
| levonorgestrel | 3.277 | 1.141 | 0.926 | 0 | 0 | 0 | 10.249 | 8.334 | 3.999 |
| levothyroxine | 24.486 | 44.506 | 33.591 | 0.001 | 0.005 | 0.002 | 0 | 0 | 0 |
| lomitapide | 0.133 | 0.066 | 0.146 | 0 | 0 | 0 | 0.005 | 0.006 | 0.007 |
| lormetazepam | 62.938 | 213.777 | 64.038 | 0.003 | 0.025 | 0.004 | 0.019 | 0.151 | 0.027 |
| lusutrombopag | 0.001 | 0.002 | 0.001 | 0 | 0 | 0 | 0 | 0 | 0 |
| mebendazole | 160.331 | 328.987 | 203.33 | 0.008 | 0.038 | 0.014 | 0.091 | 0.437 | 0.16 |
| metformin | 284370.474 | 588134.109 | 531497.227 | 14.228 | 68.76 | 36.734 | 1.423 | 6.876 | 3.673 |
| miconazole | 1597.412 | 3841.645 | 3677.486 | 0.08 | 0.449 | 0.254 | 3.169 | 17.809 | 10.078 |
| midazolam | 12.448 | 27.496 | 21.957 | 0.001 | 0.003 | 0.002 | 0.005 | 0.028 | 0.013 |
| midostaurin | 0.67 | 1.635 | 0.885 | 0 | 0 | 0 | 0.043 | 0.247 | 0.079 |

| | | | | | | | | | |
|---|---|---|---|---|---|---|---|---|---|
| montelukast | 93.002 | 192.262 | 181.328 | 0.005 | 0.022 | 0.013 | 2.105 | 10.171 | 5.671 |
| mycophenolate | 2861.163 | 7795.064 | 4948.974 | 0.143 | 0.911 | 0.342 | 1.084 | 6.904 | 2.591 |
| naphazoline | 27.852 | 59.653 | 39.388 | 0.001 | 0.007 | 0.003 | 0.004 | 0.019 | 0.007 |
| nebivolol | 379.693 | 800.234 | 716.353 | 0.019 | 0.094 | 0.05 | 0.01 | 0.051 | 0.027 |
| nilotinib | 68.128 | 98.994 | 136.487 | 0.003 | 0.012 | 0.009 | 2.303 | 7.82 | 6.374 |
| nisoldipine | | 0 | 0 | | 0 | 0 | | 0 | 0 |
| norethisterone | 21.734 | 12.249 | 19.746 | 0.001 | 0.001 | 0.001 | 2.175 | 2.864 | 2.73 |
| ofloxacin | 4962.679 | 9451.613 | 9140.622 | 0.248 | 1.105 | 0.632 | 9.55 | 42.5 | 24.298 |
| olmesartan | 7850.927 | 3569.703 | 8746.683 | 0.393 | 0.417 | 0.605 | 97.711 | 103.816 | 150.38 |
| omeprazole | 1532.085 | 3625.139 | 4458.261 | 0.077 | 0.424 | 0.308 | 0.274 | 1.514 | 1.1 |
| pantoprazole | 6914.531 | 13928.816 | 11109.131 | 0.346 | 1.628 | 0.768 | 0.508 | 2.391 | 1.127 |
| paracetamol | 170395.65 | 521775.659 | 294022.477 | 8.525 | 61.002 | 20.321 | 0.064 | 0.455 | 0.152 |
| permethrin | 457.486 | 647.233 | 624.186 | 0.023 | 0.076 | 0.043 | 114.445 | 378.346 | 215.703 |
| piperaquine | 0.439 | 2.221 | 0.658 | 0 | 0 | 0 | 0.021 | 0.245 | 0.043 |
| proglumetacin | 0.288 | 0.6 | 4.044 | 0 | 0 | 0 | 0.008 | 0.041 | 0.163 |
| propranolol | 446.849 | 1476.887 | 738.945 | 0.022 | 0.173 | 0.051 | 0.224 | 1.727 | 0.511 |
| ramipril | 2616.369 | 1111.554 | 1465.531 | 0.131 | 0.13 | 0.101 | 1.194 | 1.186 | 0.924 |
| rifaximin | 6523.591 | 12207.386 | 11468.372 | 0.326 | 1.427 | 0.793 | 131.608 | 575.479 | 319.612 |
| rilpivirine | 34.358 | 101.796 | 24.127 | 0.002 | 0.012 | 0.002 | 0.677 | 4.685 | 0.657 |
| rosuvastatin | 3652.356 | 1405.255 | 2359.806 | 0.183 | 0.164 | 0.163 | 0.678 | 0.61 | 0.605 |
| sertraline | 2461.388 | 6594.448 | 2541.236 | 0.123 | 0.771 | 0.176 | 13.101 | 82.018 | 18.685 |
| simvastatin | 4840.597 | 2161.669 | 3656.918 | 0.242 | 0.253 | 0.253 | 0.092 | 0.096 | 0.096 |
| sulfamethoxazole | 1708.905 | 740.764 | 1212.414 | 0.085 | 0.087 | 0.084 | 0.855 | 0.866 | 0.838 |
| tamsulosin | 21.509 | 53.178 | 30.299 | 0.001 | 0.006 | 0.002 | 0.003 | 0.018 | 0.006 |
| toremifene | 0.437 | 0.74 | 0.326 | 0 | 0 | 0 | 0.073 | 0.288 | 0.075 |
| trimethoprim | 8026.04 | 3440.008 | 5598.163 | 0.402 | 0.402 | 0.387 | 0.803 | 0.804 | 0.774 |
| valsartan | 18919.947 | 8692.322 | 14440.684 | 0.947 | 1.016 | 0.998 | 0.002 | 0.002 | 0.002 |
| venlafaxine | 1882.582 | 4737.363 | 2341.547 | 0.094 | 0.554 | 0.162 | 15.441 | 90.796 | 26.53 |
| verteporfin | 0.002 | 0.008 | 0.002 | 0 | 0 | 0 | 0 | 0 | 0 |
| vinflunine | 0.027 | 0.104 | 0.04 | 0 | 0 | 0 | 0.001 | 0.012 | 0.003 |

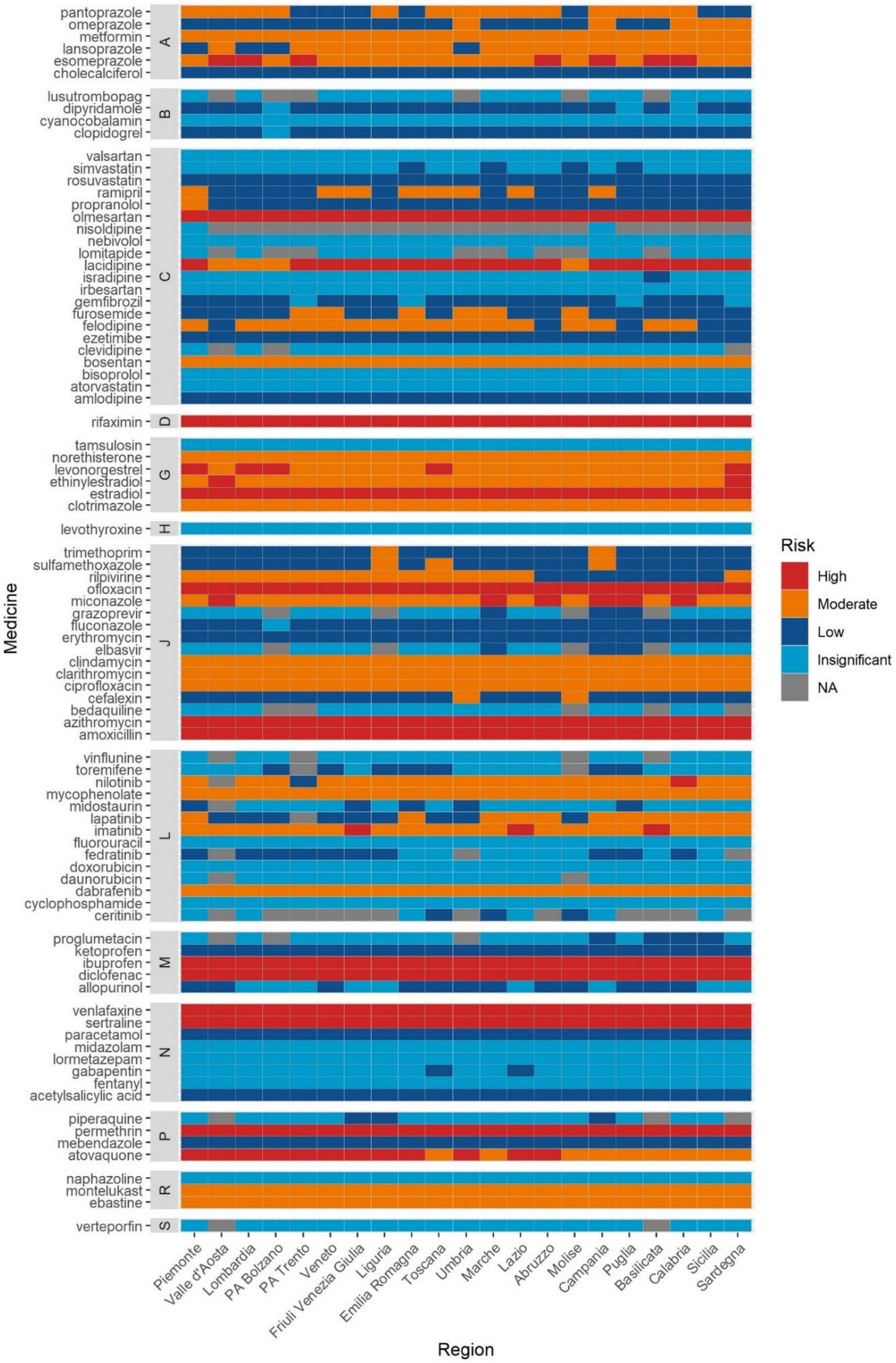


**Figure S1** – Risk level of the 90 selected medicines in 2022 according to the Italian region
