## Supplementary material for "Assessing the environmental impact of medicines in Italy using data from the Italian Medicines Agency": script_data: script_article.R

```
# Assessing the environmental impact of medicines in Italy using drug utilization data
# script of the analyses
# Giunchi V. and Fusaroli M.
# last update 20-03-2024

# libraries upload -------------------------------------------------------------
library(readxl)
library(data.table)
library(dplyr)
library(ggplot2)
ATC <- setDT(read.csv2("ATC_DiAna.csv"))

# data upload ------------------------------------------------------------------
# Italy
DU <- setDT(read_xlsx("consumption.xlsx"))
# Regions
DU_regions <- setDT(read_xlsx("consumption_reg.xlsx"))
colnames(DU_regions) <- c("Substance", "PIEMONTE", "VALLE D'AOSTA", "LOMBARDIA", "PA BOLZANO", "PA TRENTO",
                              "VENETO", "FRIULI VENEZIA GIULIA", "LIGURIA", "EMILIA ROMAGNA", "TOSCANA", "UMBRIA",
                              "MARCHE", "LAZIO", "ABRUZZO", "MOLISE", "CAMPANIA", "PUGLIA", "BASILICATA",
                              "CALABRIA", "SICILIA", "SARDEGNA")
# DU in macro-areas in DDD
DU_areas <- setDT(read_xlsx("consumption_areas.xlsx"))
colnames(DU_areas) <- c("Substance", "Centre", "North", "South")
# PNEC data
PNEC <- setDT(read_xlsx("PNEC.xlsx"))
PNEC$Substance <- tolower(PNEC$Substance)
# inhabitants data
pop <- setDT(read_xlsx("pop.xlsx"))
pop[, mean:=(POP2022+POP2023)/2, by=Regione]

# analysis 1: Italy ------------------------------------------------------------
DU[, PEC := (cons_tot_kg*10^9 *100)/(365*pop[Regione=="Italia"]$mean*200*10*100)]
DU <- left_join(DU, PNEC)
DU <- setDT(DU)
DU[, RQ := PEC/PNEC_mcg_L]
DU[RQ>=10, Risk := "High"]
DU[RQ>=1 & RQ<10, Risk := "Moderate"]
DU[RQ>=0.1 & RQ<1, Risk := "Low"]
DU[RQ<0.1, Risk := "Insignificant"]
writexl::write_xlsx(DU, "Risk analysis ITA.xlsx")

# visualization
DU_ita <- DU[, .(Substance, RQ, Risk)]
DU_ita <- left_join(DU_ita, ATC[, .(Substance, primary_code)], by = "Substance")
DU_ita <- left_join(DU_ita, ATC[, .(Substance, code, Lvl1)], by=c("Substance", "primary_code"="code"))
DU_ita <- DU_ita %>% distinct()
DU_ita[Substance == "mycophenolate"]$Lvl1 <- "L"
DU_ita[Substance == "cyanocobalamin"]$Lvl1 <- "B"
DU_ita[Substance == "cholecalciferol"]$Lvl1 <- "A"

DU_ita$Risk <- factor(DU_ita$Risk, levels = c("High","Moderate","Low","Insignificant"))
png(filename = "RQ_ita.png", width = 25, height = 30, units = "cm", res = 300)
ggplot(data = DU_ita) +
  geom_col(aes(x=(RQ+1), y=Substance, fill=Risk)) +
  scale_x_continuous(trans = "log10",breaks = c(1.1,2,11), labels = c("0.1","1","10")) +
  geom_vline(xintercept = 1.1, alpha=0.5, color="white")+
  geom_vline(xintercept = 2, alpha=0.5, color="white")+
  geom_vline(xintercept = 11, alpha=0.5, color="white") +
  scale_fill_manual(values = c("High"="firebrick3", "Moderate"="darkorange2", "Low"="dodgerblue4",
                               "Insignificant"="deepskyblue3", "NA"="gainsboro")) +
  labs(x="Risk Quotient", y="Medicine") +
  facet_grid(factor(Lvl1, levels = c("A", "B", "C", "D", "G", "H", "J", "L", "M", "N", "P", "R", "S"), ordered = TRUE)~.,
             scales = "free", space = "free", switch = "y")
dev.off()

# analysis 2: Regions ----------------------------------------------------------
DU_regions[, PEC_Piemonte := (PIEMONTE*10^9 *100)/(365*pop[Regione=="Piemonte"]$mean*200*10*100)]
DU_regions[, PEC_ValleDAosta := (`VALLE D'AOSTA`*10^9 *100)/(365*pop[Regione=="Valle d'Aosta"]$mean*200*10*100)]
DU_regions[, PEC_Lombardia := (LOMBARDIA*10^9 *100)/(365*pop[Regione=="Lombardia"]$mean*200*10*100)]
DU_regions[, PEC_Bolzano := (`PA BOLZANO`*10^9 *100)/(365*pop[Regione=="Bolzano"]$mean*200*10*100)]
DU_regions[, PEC_Trento := (`PA TRENTO`*10^9 *100)/(365*pop[Regione=="Trento"]$mean*200*10*100)]
DU_regions[, PEC_Veneto := (VENETO*10^9 *100)/(365*pop[Regione=="Veneto"]$mean*200*10*100)]
DU_regions[, PEC_Friuli := (`FRIULI VENEZIA GIULIA`*10^9 *100)/(365*pop[Regione=="Friuli Venezia Giulia"]$mean*200*10*100)]
DU_regions[, PEC_Liguria := (LIGURIA*10^9 *100)/(365*pop[Regione=="Liguria"]$mean*200*10*100)]
DU_regions[, PEC_EmiliaRomagna := (`EMILIA ROMAGNA`*10^9 *100)/(365*pop[Regione=="Emilia Romagna"]$mean*200*10*100)]
DU_regions[, PEC_Toscana := (TOSCANA*10^9 *100)/(365*pop[Regione=="Toscana"]$mean*200*10*100)]
DU_regions[, PEC_Umbria := (UMBRIA*10^9 *100)/(365*pop[Regione=="Umbria"]$mean*200*10*100)]
DU_regions[, PEC_Marche := (MARCHE*10^9 *100)/(365*pop[Regione=="Marche"]$mean*200*10*100)]
DU_regions[, PEC_Lazio := (LAZIO*10^9 *100)/(365*pop[Regione=="Lazio"]$mean*200*10*100)]
DU_regions[, PEC_Abruzzo := (ABRUZZO*10^9 *100)/(365*pop[Regione=="Abruzzo"]$mean*200*10*100)]
DU_regions[, PEC_Molise := (MOLISE*10^9 *100)/(365*pop[Regione=="Molise"]$mean*200*10*100)]
DU_regions[, PEC_Campania := (CAMPANIA*10^9 *100)/(365*pop[Regione=="Campania"]$mean*200*10*100)]
DU_regions[, PEC_Puglia := (PUGLIA*10^9 *100)/(365*pop[Regione=="Puglia"]$mean*200*10*100)]
DU_regions[, PEC_Basilicata := (BASILICATA*10^9 *100)/(365*pop[Regione=="Basilicata"]$mean*200*10*100)]
DU_regions[, PEC_Calabria := (CALABRIA*10^9 *100)/(365*pop[Regione=="Calabria"]$mean*200*10*100)]
DU_regions[, PEC_Sicilia := (SICILIA*10^9 *100)/(365*pop[Regione=="Sicilia"]$mean*200*10*100)]
DU_regions[, PEC_Sardegna := (SARDEGNA*10^9 *100)/(365*pop[Regione=="Sardegna"]$mean*200*10*100)]
DU_regions <- left_join(DU_regions, PNEC)
DU_regions <- setDT(DU_regions)
DU_regions[, RQ_Piemonte := PEC_Piemonte/PNEC_mcg_L]
DU_regions[, RQ_ValleDAosta := PEC_ValleDAosta/PNEC_mcg_L]
DU_regions[, RQ_Lombardia := PEC_Lombardia/PNEC_mcg_L]
DU_regions[, RQ_Bolzano := PEC_Bolzano/PNEC_mcg_L]
DU_regions[, RQ_Trento := PEC_Trento/PNEC_mcg_L]
DU_regions[, RQ_Veneto := PEC_Veneto/PNEC_mcg_L]
DU_regions[, RQ_Friuli := PEC_Friuli/PNEC_mcg_L]
DU_regions[, RQ_Liguria := PEC_Liguria/PNEC_mcg_L]
DU_regions[, RQ_EmiliaRomagna := PEC_EmiliaRomagna/PNEC_mcg_L]
DU_regions[, RQ_Toscana := PEC_Toscana/PNEC_mcg_L]
DU_regions[, RQ_Umbria := PEC_Umbria/PNEC_mcg_L]
DU_regions[, RQ_Marche := PEC_Marche/PNEC_mcg_L]
DU_regions[, RQ_Lazio := PEC_Lazio/PNEC_mcg_L]
DU_regions[, RQ_Abruzzo := PEC_Abruzzo/PNEC_mcg_L]
DU_regions[, RQ_Molise := PEC_Molise/PNEC_mcg_L]
DU_regions[, RQ_Campania := PEC_Campania/PNEC_mcg_L]
DU_regions[, RQ_Puglia := PEC_Puglia/PNEC_mcg_L]
DU_regions[, RQ_Basilicata := PEC_Basilicata/PNEC_mcg_L]
DU_regions[, RQ_Calabria := PEC_Calabria/PNEC_mcg_L]
DU_regions[, RQ_Sicilia := PEC_Sicilia/PNEC_mcg_L]
DU_regions[, RQ_Sardegna := PEC_Sardegna/PNEC_mcg_L]
writexl::write_xlsx(DU_reg, "Risk analysis REG.xlsx")

# visualization
DU_reg_RQ <- DU_regions[, .(Substance, RQ_Piemonte, RQ_ValleDAosta, RQ_Lombardia, RQ_Bolzano, RQ_Trento, RQ_Veneto,
                        RQ_Friuli, RQ_Liguria, RQ_EmiliaRomagna, RQ_Toscana, RQ_Umbria, RQ_Marche, RQ_Lazio,
                        RQ_Abruzzo, RQ_Molise, RQ_Campania, RQ_Puglia, RQ_Basilicata, RQ_Calabria, RQ_Sicilia,
                        RQ_Sardegna)]
DU_reg_long <- melt(DU_reg_RQ, id.vars = "Substance")
colnames(DU_reg_long) <- c("Substance", "Region", "RQ")
DU_reg_long <- setDT(DU_reg_long)
DU_reg_long[, Risk := ifelse(RQ>=10, "High", ifelse(RQ>=1, "Moderate", ifelse(RQ>=0.1, "Low", "Insignificant")))]
DU_reg_long$Risk <- factor(DU_reg_long$Risk, levels=c("High", "Moderate", "Low", "Insignificant"))
DU_reg_long$Substance <- tolower(DU_reg_long$Substance)
DU_reg_long <- left_join(DU_reg_long, ATC[, .(Substance, primary_code)], by = "Substance")
DU_reg_long <- left_join(DU_reg_long, ATC[, .(Substance, code, Lvl1)], by=c("Substance", "primary_code"="code"))
DU_reg_long <- DU_reg_long %>% distinct()
DU_reg_long[Substance == "mycophenolate"]$Lvl1 <- "L"
DU_reg_long[Substance == "cyanocobalamin"]$Lvl1 <- "B"
DU_reg_long[Substance == "cholecalciferol"]$Lvl1 <- "A"

DU_reg_long$Risk <- factor(DU_reg_long$Risk, levels = c("High", "Moderate", "Low", "Insignificant"))

png(filename = "RQ_reg.png", width = 20, height = 30, units = "cm", res = 300)
ggplot(data=DU_reg_long) +
  geom_tile(aes(x=Region, y=Substance, fill=Risk), color="gray") +
  scale_fill_manual(values = c("High"="firebrick3", "Moderate"="darkorange2", "Low"="dodgerblue4",
                               "Insignificant"="deepskyblue3", "NA"="gainsboro")) +
  scale_x_discrete(labels = c("Piemonte", "Valle d'Aosta", "Lombardia", "PA Bolzano", "PA Trento", "Veneto",
                              "Friuli Venezia Giulia", "Liguria", "Emilia Romagna", "Toscana", "Umbria",
                              "Marche", "Lazio", "Abruzzo", "Molise", "Campania", "Puglia", "Basilicata",
                              "Calabria", "Sicilia", "Sardegna")) +
  facet_grid(factor(Lvl1, levels = c("A", "B", "C", "D", "G", "H", "J", "L", "M", "N", "P", "R", "S"), ordered = TRUE)~.,
             scales = "free", space = "free", switch = "y")+
  ylab("Medicine")+
  theme(axis.text.x = element_text(angle = 45, hjust=1))
dev.off()


# Analysis 3: macro-areas ------------------------------------------------------
DU_areas[, PEC_North := (North*10^9 *100)/(365*pop[Regione=="Nord"]$mean*200*10*100)]
DU_areas[, PEC_Centre := (Centre*10^9 *100)/(365*pop[Regione=="Centro"]$mean*200*10*100)]
DU_areas[, PEC_South := (South*10^9 *100)/(365*pop[Regione=="Sud"]$mean*200*10*100)]
DU_areas <- left_join(DU_areas, PNEC)
DU_areas <- setDT(DU_areas)
DU_areas[, RQ_North := PEC_North/PNEC_mcg_L]
DU_areas[, RQ_Centre := PEC_Centre/PNEC_mcg_L]
DU_areas[, RQ_South := PEC_South/PNEC_mcg_L]
writexl::write_xlsx(DU_areas, "Risk analysis AREAS.xlsx")

# visualization
DU_areas_RQ <- DU_areas[, .(Substance, RQ_North, RQ_Centre, RQ_South)]
DU_areas_long <- melt(DU_areas_RQ, id.vars = "Substance")
colnames(DU_areas_long) <- c("Substance", "Area", "RQ")
DU_areas_long <- setDT(DU_areas_long)
DU_areas_long[, Risk := ifelse(RQ>=10, "High", ifelse(RQ>=1, "Moderate", ifelse(RQ>=0.1, "Low", "Insignificant")))]
DU_areas_long$Risk <- factor(DU_areas_long$Risk, levels=c("High", "Moderate", "Low", "Insignificant"))

DU_areas_long$Substance <- tolower(DU_areas_long$Substance)
DU_areas_long <- left_join(DU_areas_long, ATC[, .(Substance, primary_code)], by = "Substance")
DU_areas_long <- left_join(DU_areas_long, ATC[, .(Substance, code, Lvl1)], by=c("Substance", "primary_code"="code"))
DU_areas_long <- DU_areas_long %>% distinct()
DU_areas_long[Substance == "mycophenolate"]$Lvl1 <- "L"
DU_areas_long[Substance == "cyanocobalamin"]$Lvl1 <- "B"
DU_areas_long[Substance == "cholecalciferol"]$Lvl1 <- "A"

DU_areas_long$Risk <- factor(DU_areas_long$Risk, levels = c("High", "Moderate", "Low", "Insignificant"))

png(filename = "RQ_areas.png", width = 10, height = 30, units = "cm", res = 300)
ggplot(data=DU_areas_long) +
  geom_tile(aes(x=Area, y=Substance, fill=Risk), color="gray") +
  scale_fill_manual(values = c("High"="firebrick3", "Moderate"="darkorange2", "Low"="dodgerblue4",
                               "Insignificant"="deepskyblue3", "NA"="gainsboro")) +
  scale_x_discrete(labels = c("North", "Centre", "South")) +
  facet_grid(factor(Lvl1, levels = c("A", "B", "C", "D", "G", "H", "J", "L", "M", "N", "P", "R", "S"), ordered = TRUE)~.,
             scales = "free", space = "free", switch = "y")+
  ylab("Medicine")+
  theme(axis.text.x = element_text(angle = 45, hjust=1))
dev.off()
```
